# Assessing Heart Rate Variability Responses to Food Cues and Cognitive Stress: A Comparative Study of a Wearable Device and ECG

**DOI:** 10.1101/2025.10.22.25338529

**Authors:** Sirin W. Gangstad, Maika Lindkvist Jensen, Anne-Marie Raabyemagle, Akasha P. Strub, Monica Jane Emerson, Jon Davis, Anne Raben, Lena-Sophie Martis

## Abstract

Variations in heart rate variability (HRV) have been linked to emotional distress, disordered eating and exposure to food stimuli, suggesting HRV as a potential biomarker for emotional dysregulation related to maladaptive eating. With advancements in wearable technology, continuous HRV monitoring is now feasible. This study explored HRV responses to visual food cues, food consumption, and cognitive stress in healthy individuals using both a wrist-worn wearable device and a 12-lead electrocardiography (ECG), the gold standard for HRV measurements. The motivation behind the study was to investigate the feasibility of detecting a binge eating episode prior to the event, as a first step to subsequently developing a just-in-time intervention. Unlike previous studies, no conclusive food-elicited HRV responses were found with the ECG. However, the ECG did capture a significant difference in HRV from baseline compared to the cognitive stress test. The wrist-worn device’s signal quality was insufficient for a reliable comparison with the ECG. While wearable devices can continuously measure heart activity, this study highlights the susceptibility of their sensors to motion interference and indicates that signals should be interpreted with care.

## Introduction

Binge-eating disorder is the most prevalent form of eating disorder (ED), affecting millions globally with an estimated lifetime prevalence of 2% and higher (Kessler et al., 2013; Nagata et al., 2023). The disorder is characterized by episodes of consuming large amounts of food, accompanied by a sense of loss of control and often followed by feeling guilty or depressed (Berkman, 2015; NIDDK, 2021). Previous studies have found an association between disordered eating behaviors and heart rate variability (HRV), suggesting HRV can be used as a physiological proxy of self-regulation (Christensen et al. (2023); Godfrey et al. (2019).

HRV can be characterized using time- and frequency-domain metrics. Time-domain measures analyze the intervals between successive normal heartbeats given in milliseconds, commonly metricized as root mean square of successive differences between RR intervals (RMSSD) or as the standard deviation of NN intervals. Frequency-domain measures use spectral analysis of heart rate (HR) changes, metricized as the power in frequency intervals given in ms^2^; common frequency intervals include low frequencies (LF) defined as the band from 0.04–0.15 Hz, and high frequencies (HF), defined as 0.15–0.4 Hz (Shaffer & Ginsberg, 2017).

Godfrey et al. investigated the link between HRV and binge-eating severity in patients with obesity and loss-of-control eating (Godfrey et al., 2019). By recording HRV at rest and during a mental stressor in a controlled laboratory environment, they found that a lower time-domain HRV during rest was linked to more severe self-reported loss of control. In the frequency domain, there was an increased LF and decreased HF in the stressed conditions compared to the resting condition. Furthermore, the magnitude of these frequency-domain changes was associated with more severe self-reported overeating.

Chang et al. studied the influence of body mass index (BMI), self-reported disordered eating behavior, and sex on the HRV response to food exposure in healthy adults (Chang et al., 2021). The participants completed questionnaires assessing their disordered eating symptoms, and HRV was recorded while the participants were exposed to visual stimuli of high-energy food, neutral cues and negative emotional stimuli in a controlled laboratory setting. They found an increase in frequency-domain HRV reactivity to food signals that was modulated by BMI. The authors suggest that BMI might influence the interaction between sympathetic activity and food stimuli, but that the link between this interaction and diet control still warrants further investigation.

The studies mentioned above captured the HRV by recording standard clinical 12-lead ECG or other chest-worn HRV assessment equipment. Although clinical ECG signals display a good signal-to-noise ratio when recorded properly in a controlled laboratory setting, this monitoring method has the disadvantage of not capturing transient and rare events manifesting outside the clinic. ECG is therefore not suited to capture momentary HRV responses in a free-living environment. However, the advancements of photoplethysmography (PPG) sensors incorporated into wearable digital health technologies (DHTs) now allows for estimating the pulse rate and pulse rate variability (PRV) by recording changes in blood volume in the tissue monitored by the PPG sensor.

The technological development of PPG sensors has motivated research into the association between changes in PRV prior to binge eating episodes as they unfold in real life. Juarascio et al. utilized a wrist-worn DHT to distinguish PRV preceding emotional eating episodes to control time periods in adults with clinically significant emotional eating behaviors (Juarascio et al., 2020). Furthermore, Ralph-Nearman et al. examined PR, electrodermal activity (EDA) and peripheral skin temperature (PST) in six individuals diagnosed with an eating disorder (Ralph-Nearman et al., 2024). They were able to distinguish between time windows immediately preceding an eating disorder-related event and control windows. Both studies argue that the results show promise for wrist-worn devices to detect an imminent disordered eating episode allowing for a just-in-time intervention (Juarascio et al., 2018).

However, several studies have compared ECG-measured HRV recorded near the heart and PPG-measured PRV recorded at the wrist and noted discrepancies under certain conditions. At the physiological level, the discrepancies could be explained by the hemodynamics of arterial pulse wave propagation, blood content, blood pressure and respiratory frequency (Constant et al., 1999). The discrepancies can be influenced by experimental conditions such as ambient temperature, body position, exercise, and stress exposure (Charlot et al., 2009; Giardino et al., 2002; Shin, 2016). Furthermore, as the HRV and PRV are not measured using the same sensor technology, experimental conditions affect the signal quality of the two measurement modalities differently at the sensor level. Combined, the differentiated effect on the measurement modalities affects the accuracy of the PRV compared to the HRV.

Given the observed discrepancies, it is necessary to demonstrate that the link between HRV and eating disorder is equally valid for PRV. Therefore, in this study, PRV and HRV were simultaneously recorded from healthy volunteers exposed to food cues in a fasted and full condition, during meal consumption and a mild stress test as control condition.

The main research objective was to investigate whether there is a significant change in HRV in response to visual food cues, a meal and cognitive stress as measured by reference ECG. Furthermore, if such a change was found, to assert if the same change could be observed for PRV as measured by the wrist-worn device.

## Methods

### Procedure summary

The study participants were healthy volunteers. The data collection consisted of a two-week period of free-living, at-home monitoring using the Empatica EmbracePlus, a CE-certified and medical-grade device (Empatica Inc.), worn on the non-dominant wrist, followed by a test day in a controlled laboratory setting. On the test day, data was collected using the Empatica wristband and a 12-lead ECG (Seer 12, GE HealthCare). Pre-specified endpoints were computed both on the HRV derived from the ECG and using PRV derived from the Empatica EmbracePlus. A subset of the endpoints were based on the electrodermal activity (EDA) signal recorded by the wrist worn device.

### Ethical approval and informed consent

The study was performed in line with the principles of the declaration of Helsinki and was approved by the Ethics Committee of the Capital Region of Denmark (H-23025030).

### Participants

Participants were healthy volunteers recruited through web portals, social media, local newspapers and flyers. Additionally, individuals on a waiting list in a local database or previous study participants who had expressed interest in future studies were contacted. The recruitment targeted individuals aged 25-55 years with a body mass index (BMI) ≥18.5 kg/m^2^. At least 40% of participants were required to have a BMI of 18.5-29.9 kg/m^2^, while another 40% needed a BMI of ≥30.0 kg/m^2^. The study aimed to include 50 participants, anticipating that 45 would complete the study.

Exclusion criteria were recent treatment with investigational drugs, physical or mental inability to comply with the study procedures, lack of access to phone or internet, known or suspected eating disorders, recent surgical or medical treatment for obesity, diabetes mellitus, cardiovascular disease, significant liver disease or uncontrolled thyroid disease, certain medications, active malignancy, certain chronic diseases, psychiatric illness, pregnancy or nursing, infrequent breakfast consumption, tobacco use, substance abuse, night or shift work, and intensive physical training.

All participants signed informed consent forms after receiving written and oral information before initiating any study-related activities.

### Data collection

#### At-home monitoring

After signing the informed consent, participants were equipped with the Empatica EmbracePlus wearable device, which they wore for two weeks to collect free-living baseline PRV measurements. Participants were instructed on proper usage, including charging and data transfer procedures. The at-home data was used to better understand the inter- and intra-participant variability and the signal-to-noise ratio of the recorded signal.

#### In-lab test day

On the morning of the test day, participants arrived at the clinic after fasting for at least 10 hours to increase their responsiveness to food. Upon arrival, the 12-lead ECG was mounted to record HRV serving as a reference for the wearable device-measured PRV. Then followed a baseline recording where the participants were instructed to rest quietly for 45 minutes and refrain from excessive movement. The first 10 and last five minutes of the baseline recording were excluded from analysis to ensure a stable signal. To be able to account for varying levels of appetite, ghrelin measurements were taken from plasma, and participants were asked to provide subjective assessments of hunger and satiety via visual analogue scale (VAS) scores (Flint et al., 2000).

Following a 20-minute break to reduce potential effects on HRV from blood draw for ghrelin assessment, participants were exposed to a test block presenting images of either food-related cues or neutral cues. Each cue block contained 49 images and lasted 5 minutes.

Food and neutral cues were designed to match visual complexity. Next, participants were presented with a meal, which they were instructed to look and smell for 5 minutes before consuming it. Data recorded while looking at, smelling and consuming the meal was not analyzed. After the meal and a short break, hunger and satiety were again assessed through both methods mentioned above. Following a new baseline measurement, the test block containing images with and without food cues was repeated.

Finally, the test day concluded with a Stroop test to evaluate the physiological response to cognitive stress, which also acted as positive control in regard to HRV response. The test day is graphically illustrated in Figure 1.

**Figure 1.**
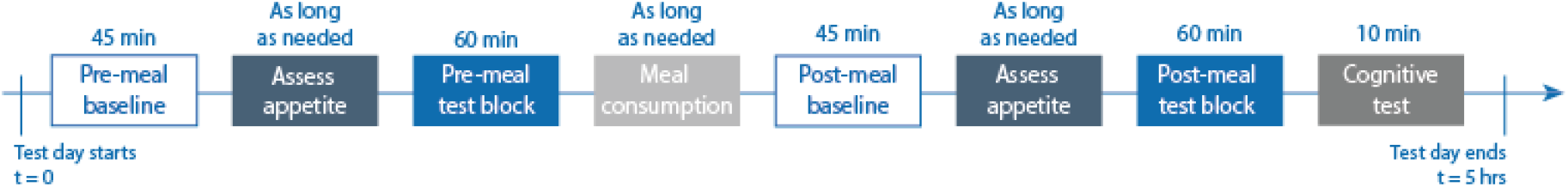
Timeline of the test day: Illustration of the activities recorded by the wrist-worn device and 12-lead ECG reference. The test day started after the participant arrived fasted at site, was briefed about the upcoming activities, had the 12-lead ECG mounted and the wearable device was confirmed to be worn correctly and recording. The “Assess appetite” block represents both the objective and subjective measurements of hunger and satiety, and the “Meal consumption” block represents both the looking at and smelling the meal and consuming the meal.

### Data pre-processing

QRS timestamps – representing the timing of each heartbeat – were extracted from the ECG using proprietary software (CardioDay version 2.6, GE HealthCare). HRV was computed both in the time domain and in the frequency domain using Python (version 3.11). The time domain measure used was the root mean square of successive differences (RMSSD) between normal heartbeats, computed in consecutive, non-overlapping 5-minute windows. The RMSSD is an estimate of the vagally mediated changes in HRV and largely reflects the parasympathetic activity (Peschel et al., 2016; Shaffer et al., 2014). The frequency domain measures used were high frequency (HF) HRV, reflecting parasympathetic activity (Peschel et al., 2016), and the ratio between high and low frequencies LF/HF HRV, reflecting the ratio between the activity of the sympathetic nervous system (SNS) and the parasympathetic nervous system (PNS) (Shaffer & Ginsberg, 2017; Shaffer et al., 2014). Both frequency measures were evaluated in consecutive, non-overlapping 5-minute windows. The frequency content was computed using the Python package NeuroKit2 (version 0.2.11), where the low and high frequency intervals were defined as 0.04 Hz - 0.15 Hz and 0.15 Hz – 0.4 Hz respectively (Pham et al., 2021).

The processing pipeline used to calculate the HRV from the ECG data was also used to calculate the PRV from the data recorded by the Empatica EmbracePlus. However, instead of the passing QRS timestamps from the ECG as input to the pipeline, the timestamps of the peaks in the blood volume pulse (BVP) signal from the wearable were used. As the BVP signal is susceptible to motion artefacts, peaks resulting from or influenced by motion artefacts were removed prior to extracting the BVP peak timestamps. Identification of motion contaminated peaks was done by exploiting one of the post-processed signals offered by the device manufacturer. This signal contains a version of PRV computed in consecutive, non-overlapping 1-minute windows. However, a PRV estimate is only provided for the 1-minute windows in which *all* BVP peaks are considered clean. By masking the continuous BVP signal with a representation of the clean 1-minute windows, windows containing contaminated BVP signal were discarded prior to PRV analysis. The PRV estimates were computed based on the systolic peak timestamps for the remaining clean peaks in 5-minute epochs. HRV and PRV estimates computed from 5-minute epochs recorded during the same type of exposure and conditions were averaged. EDA metrics based on data from the wearable were similarly computed in 5-minute epochs and averaged across the same exposure type and condition.

### Statistical analysis

The statistical analysis was performed using a linear mixed model to account for within-participant variation, as HRV is highly individual. The mixed model incorporated fasting ghrelin levels, sex, BMI, hip-to-waist ratio, waist circumference, and age as fixed effect covariates, and participant as random effect. Statistical analyses were conducted using R (version 4.3.1) and the library lme4 (version 1.1-36).

The primary endpoint of the study was to assess the difference in HRV and PRV as measured by the RMSSD between the pre-meal baseline and the pre-meal food cue exposure.

The secondary endpoints were the difference in HRV and PRV as measured by the RMSSD and in EDA between the following conditions:

1. Pre-meal baseline and pre-meal food cue exposures (for EDA)
2. Pre-meal baseline and food consumption
3. Pre-meal neutral cue exposures and pre-meal food cue exposures
4. Pre-meal food cue exposure and post-meal food cue exposure
5. Post-meal baseline and stress exposure

The exploratory endpoints focused on HRV as defined by the frequency component. These endpoints consisted of the difference in the HF band and the LF/HF ratio between the same five conditions listed above.

## Results

Forty-seven (47) participants were recruited to the study, two of whom withdrew consent after initiation. The baseline characteristics of the final cohort are shown in **Error! Reference source not found**..

The HRVestimates for each condition are shown in Figure 2 in the main text and are listed in

**Table 1.** Baseline characteristics of all participants included in the analysis. BMI: Body mass index.

|  | **Total (N = 45)**<br>**Mean ± SD** |
| --- | --- |
| Age, years | 37.8 ± 11.2 |
| Sex (male), n (%) | 14, (31.1%) |
| BMI, kg/m<sup>2</sup> | 26.5 ± 6.3 |
| Waist circumference, cm | 88.04 ± 15.6 |
| Waist-to-hip ratio | 0.8 ± 0.1 |
| Fasting ghrelin, pg/mL | 1277 ± 598 |

**Figure 2.**
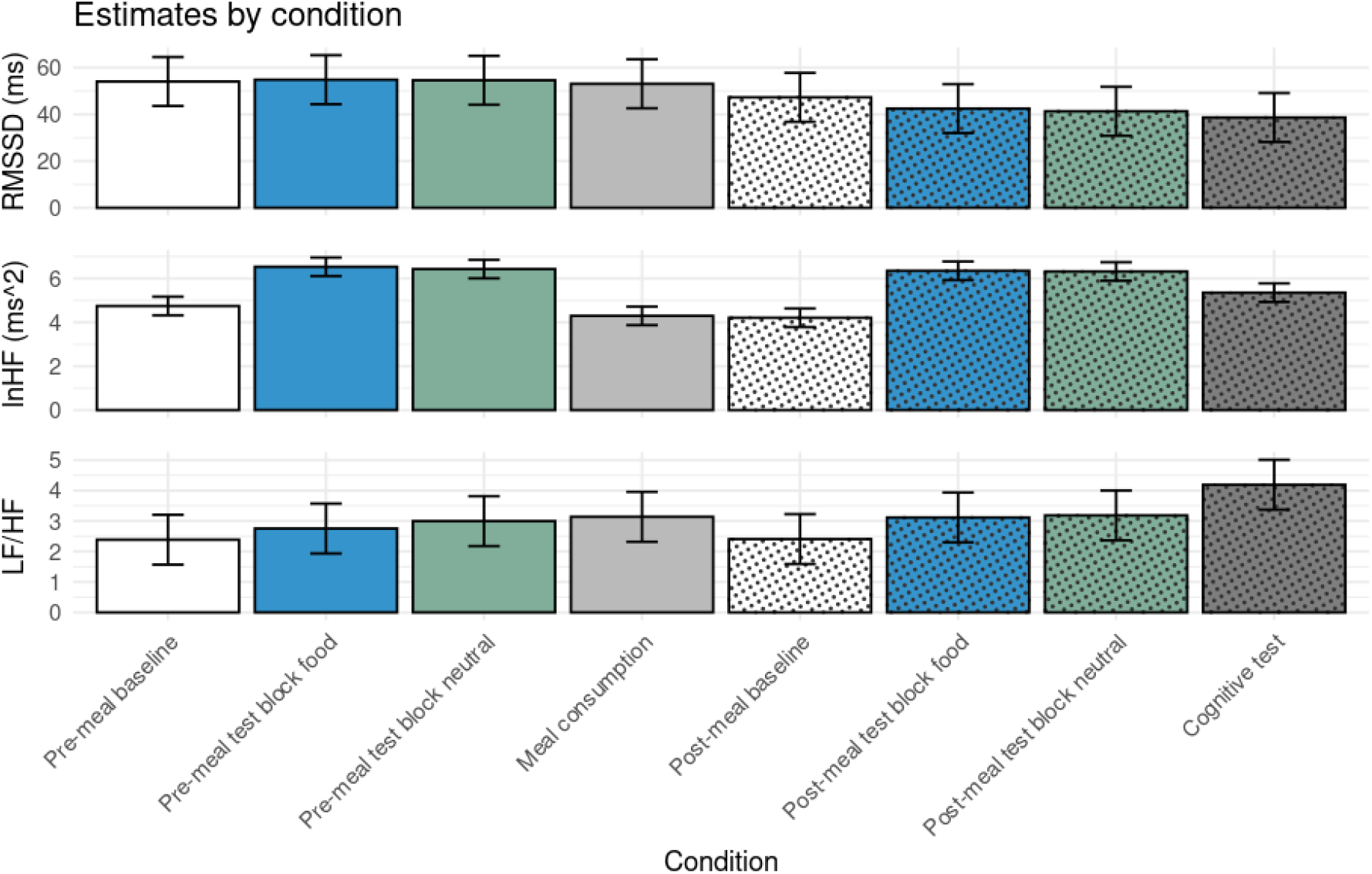
Estimated least-squares means with 95% confidence intervals for heart rate variability per experimental condition as measured by electrocardiogram. The heart rate variability is computed as root mean square of successive differences (top), high frequency component (middle) and the ratio between the low frequency to the high frequency components of the signal (bottom).

**Error! Reference source not found**., **Error! Reference source not found**. and **Error! Reference source not found**. in the Supplementary Information. No statistical effects of the HRV endpoints as measured by the ECG were observed, except for three of the exploratory endpoints:

- the difference in HF between pre-meal baseline and pre-meal food cue exposures,
- the difference in HF between post-meal baseline and stress exposure,
- the difference in LF/HF between post-meal baseline and stress exposure.

The significant difference in HRV characterized by the HF component was significant not only between the pre-meal baseline and food cues, but also between pre-meal baseline and neutral cues (post-hoc analysis). There was no significant difference between pre-meal food cues and pre-meal neutral cues.

After cleaning the BVP signal recorded by the wrist-worn device, there was insufficient data to compute the PRV related endpoints as pre-specified in the statistical analysis plan. These are therefore not reported. Analysis of the EDA signal resulted in EDA values outside of physiologically plausible ranges.

As a post-hoc analysis, the HR changes during the test day were qualitatively assessed by plotting spectrograms of the ECG overlaid by the timestamps of each test day activity. For most participants, the only conditions that clearly affected HR were drawing blood samples, the cognitive stress test, and breaking the fast. The latter appeared as a clear shift in heart activity from the hours spent while being fasted as compared to the hours after being given a meal. Although there was a substantial amount of missing data from the wrist-worn device, the data that did remain after cleaning echoed the same pattern as the ECG.

## Discussion

The aim of this study was to examine changes in HRV in healthy adults when exposed to images containing food cues and neutral cues, a meal and a cognitive stress test.

Furthermore, the study aimed to investigate whether the same link could be observed when characterizing the heart activity using PRV as opposed to HRV.

Similarly to Chang et al, there were no significant changes in HRV RMSSD between any of the pre-specified conditions as measured by the reference ECG (Chang et al., 2021). It remains unknown whether this is due to the food images not evoking a physiological response in the participants or because the RMSSD is not a sensitive enough measure.

Bourdillon et al. has previously demonstrated that RMSSD and the standard deviation of the normal to normal inter beat intervals are more sensitive to signal artefacts in healthy athletes compared to frequency-based HRV metrics (Bourdillon et al., 2022).

There was an increase in HRV HF from pre-meal baseline to pre-meal food-cue exposure. However, a significant increase in HRV HF was also observed between the pre-meal baseline and the pre-meal neutral-cue exposure, indicating that the HRV response was likely a reaction to the task of observing images and not specifically elicited by the food-cues. This result aligns with previous research finding increased parasympathetic activity when adapting to a changing environment (Christensen et al., 2023; Schmalbach et al., 2021), and thus should be considered in future study designs.

Both HRV HF and HRV LF/HF ratio showed a significant increase during the cognitive stress test compared to the post-meal baseline period. The increase in HRV HF was unexpected, as it is hypothesized to represent the parasympathetic “rest and digest” mode. The increase in LF/HF is in line with previous research suggesting that an increased LF/HF indicates a stronger SNS response relative to the PNS activity as seen in “fight-or-flight” mode (Delaney & Brodie, 2000; Kim et al., 2018). Although the observed changes partly conflict with the current understanding of the association between the nervous system and frequency-based HRV metrics, it does support the concept of monitoring the heart to detect responses to stressful situations.

It was not possible to examine whether the responses of the heart activity could also be seen when measured by the PRV. This was due to the conservative cleaning approach used for the signal recorded by the wrist-worn device. Although this approach worked well for the ECG data, it did not leave enough clean data to compute the pre-specified endpoints based on the wearable data. Potential improvements to the approach include (1) a more granular representation of which sections of the BVP signal should be discarded due to motion artefacts. In the current study, a 1-minute window would be discarded if only a single peak in that window was contaminated, leading to an unnecessary exclution of the remaining good peaks. Secondly (2), our suggestion is to impose less strict requirements of how many clean, consecutive systolic peaks are required to estimate the PRV or (3) using a longer analysis window than 5 minutes to optimize the probability of recording enough clean systolic peaks to reliably estimate the PRV. As an example, when Ralph-Nearman et al. examined pulse rate, EDA and peripheral skin temperature prior to eating disorder-related episodes, they analyzed 20-minute windows by extracting features that required minimal signal cleaning (Ralph-Nearman et al., 2024). Juarascio et al. used 30-minute windows when analyzing the PRV prior to binge eating episodes (Juarascio et al., 2020).

Furthermore, they used interpolation to handle periods with insufficient data when appropriate. It is worth noting that the consequence of using a longer analysis window is a reduced temporal resolution of the output. Consequently, short-lived responses to stressors which can be recorded using high fidelity laboratory equipment, such as ECG, might not be possible to detect when using alternative measurement modalities, such as wearable sensors, requiring longer analysis windows.

A limitation of the current study is that, although the overall aim of the research project was to assess the feasibility of detecting HRV and PRV changes in the context of disordered eating, the study was conducted on healthy volunteers. As seen in Figure 2, there was no difference between any of the responses to food-cue images compared to neutral-cue images, proving the tested population had no distinctly negative emotional responses to food-cue images. To further bridge the gap between research made using laboratory reference devices and experiments using novel devices, future research is needed on the intended patient population. Furthermore, user research in patients with Binge Eating Disorder is needed to ensure that a just-in-time intervention product is patient-centric, and catches the buildup at the right time, with the right intervention.

## Conclusion

In conclusion, this study confirmed that frequency-based metrics of HRV can be used to measure stress-related responses in healthy individuals. However, to measure potentially negative or strong emotional reactions to food or food cue exposures, a study on the intended patient population (individuals with a maladaptive emotional relationship to food) needs to be conducted. Additionally, it was confirmed that photoplethysmography and electrodermal activity signals from wearables are highly sensitive to motion artefacts.

Therefore, further considerations are required regarding cleaning and analysis of the signal to allow the application of wearables for just-in-time interventions. Lastly, qualitative research is needed to further understand the underlying mechanisms and triggers of binge eating events and extract the best timing and type of intervention to benefit this patient population.

## Declarations

### Conflict-of-Interest Statement

SWG, MLJ, APS, MJE, JD, AR and LSM are employees of Novo Nordisk A/S. SWG, MLJ, APS, AR and LSM are shareholders on Novo Nordisk A/S. AR has received honorariums from the International Sweeteners Association, Nestlé and Unilever and research funds from Novo Nordisk A/S.

### Authors’ contribution

LSM, JD, MJE, AR and AMR contributed to the study conception and design. Material preparation and data collection were performed by LSM, AR and AMR. Data analysis was performed by SWG, APS, MLJ and MJE. The first draft of the manuscript was written by SWG and all authors commented on previous versions of the manuscript. All authors read and approved the final manuscript.

## Acknowledgements

The authors would like to thank Clare Cox, Lisa Olsen, Ana Calduch Arques, Marianne Willert, Robert Koivula, Ramneek Gupta, Oona Dierickx, Daniel Brunicardi Timmermann, and Sofie Skov Frost for their support and contributions.

## Funding

This study was funded by Novo Nordisk A/S, Denmark.

## Data availability

The data that support the findings of this study are not available in order to protect study participant privacy.

## Notes

### Competing Interest Statement

The authors have declared no competing interest.

### Author Declarations

The Danish Ethics committee/IRB of the Capital Region gave ethical approval for this work (H-23025030).

## References

Berkman, N. B. KA; Peat, CM. (2015). Management and Outcomes of Binge-Eating Disorder. NIH. https://www.ncbi.nlm.nih.gov/books/NBK338301/table/introduction.t1/

Bourdillon, N., Yazdani, S., Vesin, J.-M., Schmitt, L., & Millet, G. (2022). RMSSD Is More Sensitive to Artifacts Than Frequency-Domain Parameters: Implication in Athletes’ Monitoring. Journal of Sports Science and Medicine, 21, 260–266. 10.52082/jssm.2022.260

Chang, J.-C., Huang, W.-L., Liu, C.-Y., Tseng, M. M.-C., Yang, C. C. H., & Kuo, T. B. J. (2021). Heart Rate Variability Reactivity to Food Image Stimuli is Associated with Body Mass Index. Applied Psychophysiology and Biofeedback, 46(3), 271–277. 10.1007/s10484-021-09514-2

Charlot, K., Cornolo, J., Brugniaux, J. V., Richalet, J. P., & Pichon, A. (2009). Interchangeability between heart rate and photoplethysmography variabilities during sympathetic stimulations. Physiological Measurement, 30(12), 1357. 10.1088/0967-3334/30/12/005

Christensen, K. A., Feeling, N. R., & Rienecke, R. D. (2023). Meta-Analysis and Systematic Review of Resting-State High-Frequency Heart Rate Variability in Binge-Eating Disorder. 37(1), 50–63. 10.1027/0269-8803/a000307

Constant, I., Laude, D., Murat, I., & Elghozi, J.-L. (1999). Pulse rate variability is not a surrogate for heart rate variability. Clinical science (London, England : 1979), 97, 391–397. 10.1042/CS19990062

Delaney, J. P. A., & Brodie, D. A. (2000). Effects of Short-Term Psychological Stress on the Time and Frequency Domains of Heart-Rate Variability. Perceptual and Motor Skills, 91(2), 515–524. 10.2466/pms.2000.91.2.515

Flint, A., Raben, A., Blundell, J. E., & Astrup, A. (2000). Reproducibility, power and validity of visual analogue scales in assessment of appetite sensations in single test meal studies. International Journal of Obesity, 24(1), 38–48. 10.1038/sj.ijo.0801083

Giardino, N. D., Lehrer, P. M., & Edelberg, R. (2002). Comparison of finger plethysmograph to ECG in the measurement of heart rate variability. Psychophysiology, 39(2), 246–253. 10.1111/1469-8986.3920246

Godfrey, K. M., Juarascio, A., Manasse, S., Minassian, A., Risbrough, V., & Afari, N. (2019). Heart rate variability and emotion regulation among individuals with obesity and loss of control eating. Physiology & Behavior, 199, 73–78. 10.1016/j.physbeh.2018.11.009

Juarascio, A. S., Crochiere, R. J., Tapera, T. M., Palermo, M., & Zhang, F. (2020). Momentary changes in heart rate variability can detect risk for emotional eating episodes. Appetite, 152(1095-8304 (Electronic)).

Juarascio, A. S., Parker, M. N., Lagacey, M. A., & Godfrey, K. M. (2018). Just-in-time adaptive interventions: A novel approach for enhancing skill utilization and acquisition in cognitive behavioral therapy for eating disorders. International Journal of Eating Disorders, 51(8), 826–830. 10.1002/eat.22924

Kessler, R. C., Berglund, P. A., Chiu, W. T., Deitz, A. C., Hudson, J. I., Shahly, V., Aguilar-Gaxiola, S., Alonso, J., Angermeyer, M. C., Benjet, C., Bruffaerts, R., de Girolamo, G., de Graaf, R., Maria Haro, J., Kovess-Masfety, V., O’Neill, S., Posada-Villa, J., Sasu, C., Scott, K., … Xavier, M. (2013). The Prevalence and Correlates of Binge Eating Disorder in the World Health Organization World Mental Health Surveys. Biological Psychiatry, 73(9), 904–914. 10.1016/j.biopsych.2012.11.020

Kim, H. G., Cheon, E. J., Bai, D. S., Lee, Y. H., & Koo, B. H. (2018). Stress and Heart Rate Variability: A Meta-Analysis and Review of the Literature. (1738-3684 (Print)).

Nagata, J. M., Smith-Russack, Z., Paul, A., Saldana, G. A., Shao, I. Y., Al-Shoaibi, A. A. A., Chaphekar, A. V., Downey, A. E., He, J., Murray, S. B., Baker, F. C., & Ganson, K. T. (2023). The social epidemiology of binge-eating disorder and behaviors in early adolescents. Journal of Eating Disorders, 11(1), 182. 10.1186/s40337-023-00904-x

Niddk, N. (2021). Binge Eating Disorder - Definition and Facts. Retrieved 31-05-2025, from https://www.niddk.nih.gov/health-information/weight-management/binge-eating-disorder/definition-facts

Peschel, S. K. V., Feeling, N. R., Vögele, C., Kaess, M., Thayer, J. F., & Koenig, J. (2016). A systematic review on heart rate variability in Bulimia Nervosa. Neuroscience & Biobehavioral Reviews, 63, 78–97. 10.1016/j.neubiorev.2016.01.012

Pham, T., Lau, Z. J., Chen, S. H. A., & Makowski, D. (2021). Heart Rate Variability in Psychology: A Review of HRV Indices and an Analysis Tutorial. Sensors, 21(12), 3998. https://www.mdpi.com/1424-8220/21/12/3998

Ralph-Nearman, C., Sandoval-Araujo, L. E., Karem, A., Cusack, C. E., Glatt, S., Hooper, M. A., Rodriguez Pena, C., Cohen, D., Allen, S., Cash, E. D., Welch, K., & Levinson, C. A. (2024). Using machine learning with passive wearable sensors to pilot the detection of eating disorder behaviors in everyday life. Psychological Medicine, 54(6), 1084–1090. 10.1017/S003329172300288X

Schmalbach, I., Herhaus, B., Pässler, S., Runst, S., Berth, H., Wolff, S., Schmalbach, B., & Petrowski, K. (2021). Autonomic Nervous System Response to Psychosocial Stress in Anorexia Nervosa: A Cross-Sectional and Controlled Study [Original Research]. Frontiers in Psychology, 12. https://www.frontiersin.org/journals/psychology/articles/10.3389/fpsyg.2021.649848

Shaffer, F., & Ginsberg, J. P. (2017). An Overview of Heart Rate Variability Metrics and Norms. Frontiers in public health, 5(2296-2565 (Print)).

Shaffer, F., McCraty, R., & Zerr, C. L. (2014). A healthy heart is not a metronome: an integrative review of the heart’s anatomy and heart rate variability [Review]. Frontiers in Psychology, 5. 10.3389/fpsyg.2014.01040

Shin, H. (2016). Ambient temperature effect on pulse rate variability as an alternative to heart rate variability in young adult. Journal of clinical monitoring and computing, 30(6), 939–948.

