## Supplementory Information for "Assessing Heart Rate Variability Responses to Food Cues and Cognitive Stress: A Comparative Study of a Wearable Device and ECG"

### Supplementary Information

#### Methods

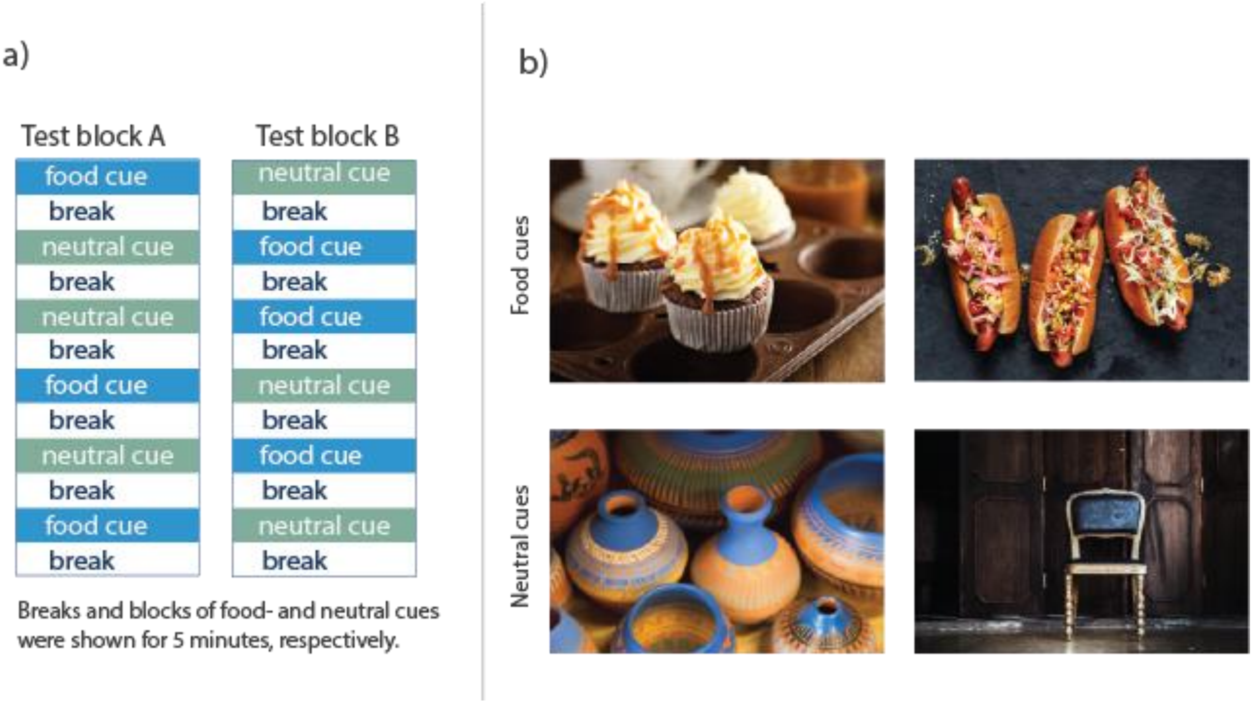


**Figure 1 -** a) Structure of test blocks**. The participants were exposed to two test blocks during the test day; one while fasted and one following a meal. The ordering of cue type (food vs neutral) within the test block differed between the pre-meal and post-meal test blocks. Participants were randomly assigned to being exposed to one of the test blocks (either block A or B) prior to the meal and the other (block B or A) after the meal.** b) Representative examples **of visual cues shown to the participants during the test blocks. Each 5-min section of either food cues or neutral cues consisted of a series of pictures similar to the examples shown in this figure.**

#### Results

| Endpoint | Estimated difference RMSSD | Lower CL | Upper CL | p-value |
|---|---|---|---|---|
| Pre-meal baseline and pre-meal food cue exposures | -0.774 | -13.724 | 12.175 | 1.000 |
| Pre-meal baseline and food consumption | -0.983 | -13.933 | 11.967 | 1.000 |

| | | | | |
|---|---|---|---|---|
| Pre-meal neutral cue exposures and pre-meal food cue exposures | 0.244 | -12.705 | 13.194 | 1.000 |
| Pre-meal food cue exposure and post-meal food cue exposure | -12.365 | -25.315 | 0.584 | 0.077 |
| Post-meal baseline and stress exposure | -8.620 | -21.570 | 4.330 | 0.561 |

**Table 1 - Estimated differences in HRV, metricized as root mean square of successive differences (RMSSD), between experimental conditions as measured by ECG.**

| Endpoint | Estimated difference LF/HF | Lower CL | Upper CL | p-value |
|---|---|---|---|---|
| Pre-meal baseline and pre-meal food cue exposures | -0.368 | -1.263 | 0.528 | 0.972 |
| Pre-meal baseline and food consumption | 0.752 | -0.143 | 1.647 | 0.201 |
| Pre-meal neutral cue exposures and pre-meal food cue exposures | -0.242 | -1.137 | 0.653 | 0.999 |
| Pre-meal food cue exposure and post-meal food cue exposure | 0.362 | -0.533 | 1.257 | 0.975 |
| Post-meal baseline and stress exposure | 1.784 | 0.889 | 2.679 | 0.000 |

**Table 2 - Estimated differences in HRV, metricized as the ratio between the low frequency and high frequency content of the HR (LF/HF), between experimental conditions as measured by ECG.**

| Endpoint | Estimated difference ln(HF) | Lower CL | Upper CL | p-value |
|---|---|---|---|---|
| Pre-meal baseline and pre-meal food cue exposures | -1.788 | -2.380 | -1.196 | 0.000 |

| | | | | |
|---|---|---|---|---|
| Pre-meal baseline and food consumption | -0.045 | -1.042 | 0.142 | 0.345 |
| Pre-meal neutral cue exposures and pre-meal food cue exposures | 0.104 | -0.488 | 0.696 | 1.000 |
| Pre-meal food cue exposure and post-meal food cue exposure | -0.183 | -0.775 | 0.409 | 0.997 |
| Post-meal baseline and stress exposure | 1.147 | 0.556 | 1.739 | 0.000 |

**Table 3 - Estimated differences in HRV, metricized as the high frequency content of the HR (HF). The values for HF are presented as the natural logarithm of HF for better interpretability.**
